# Awareness and use of the “cognitive enhancer” prescription drug modafinil in medical students

**DOI:** 10.1101/2024.08.08.24311625

**Authors:** Tatiana V Novoselova, Nyree Myatt, Esther Murray, Maryam Malekigorji, Lesley G Robson

## Abstract

**Objective:** The use of smart drugs, including modafinil, in high-pressure situations has gathered increasing attention. However, there is a lack of studies exploring their use among medical students. To investigate awareness, usage, and opinions regarding modafinil among medical students to inform student support services.

**Design:** Approximately two thousand medical students (Years 1-5) were invited to participate in an anonymous survey comprising two sections: awareness/use of modafinil and opinions on its usage. The survey collected no demographic data and ensured participants’ anonymity.

**Methods:** Online 7-minute survey using MicrosoftForms with data collection for 14 weeks.

**Results:** The survey had a low response rate. Most respondents were familiar with smart drugs, learning about them from their friends and the media. Many participants (44%) reported using modafinil to enhance attention, focus, productivity, and exam performance. Users generally found the drug effective, with some noting long-lasting effects. However, some users experienced negative effects. Analysis of opinions revealed that respondents mostly did not consider modafinil as cheating and did not feel pressured to use it if others were.

**Conclusion:** The report provides a preliminary insight into the awareness and use of modafinil in medical students The survey’s low response rate highlights the challenges of investigating drug use related topics through surveys, suggesting other methods should be employed. Nonetheless, the study underscores the need for comprehensive, professionally curated advice and policies aimed at students support to mitigate the risks.

## Introduction

Smart drugs (cognitive enhancers, or nootropics) are medications claimed to improve cognitive performance. One such drug is modafinil (sold under the brand names Modalert^®^, Provigil^®^), a non-amphetamine stimulant licensed for narcolepsy – a condition where patients struggle to stay awake and suffer from sudden “sleep attacks” (Broughton et al., 1997). It is also prescribed for shift-worker sleep disorder and sleep apnoea syndrome (Broughton et al., 1997; Hart et al., 2006). Modafinil is prescribed “off-label” for other medical conditions such as multiple sclerosis, Parkinson’s disease, chronic fatigue syndrome, depression, and attention-deficit/hyperactivity disorder, accounting for up to 89% of all prescriptions (Penaloza et al., 2013).

In healthy individuals, taking modafinil can result in an extended period of wakefulness (up to 12 hours), which some may find advantageous, particularly when working towards a pressing deadline (Kim, 2012; Bruhl et al., 2019). Numerous studies show variable degrees of increase across different modalities of cognitive performance, such as learning and memory consolidation, as well as perceived improvement of cognitive abilities (Gileen et al., 2014; Finke et al., 2010; Repantis et al., 2010; Teodorini et al., 2020). However, recent studies question the efficacy of smart drugs in terms of overall results of the cognitive effort (Bowman et al., 2023; Puyvelde et al., 2022).

The mechanisms of modafinil’s action are complex, not fully understood, and little is known about its effects during brain development. However, it is known that modafinil targets multiple pathways in the brain that promote wakefulness, alertness, and cognition (reviewed in Minzenberg and Carter, 2007). Modafinil is considered well-tolerated when taken as prescribed; however, it can cause side effects in some individuals (FDA, 2010). The most frequent side effects are headache, nausea, insomnia, anxiety, dizziness, and dry mouth. Less common side effects include chest pain or discomfort, heart palpitations, diarrhoea or abdominal pain, back pain, mood changes, severe skin reactions (Ballon et al., 2006), and very rare cases of psychosis (Wu et al., 2008; Yasui-Furukori et al., 2009).

Modafinil is also known to interact with many drugs, and some of these interactions can be highly dangerous. For instance, it can decrease the effectiveness of contraceptive pills, antihypertensive drugs, benzodiazepines, and warfarin while increasing the risks of dangerous side effects of other drugs, for example, antidepressants (selective serotonin reuptake inhibitors) (Mayo Clinic). Furthermore, the long-term side effects of modafinil use are largely unknown. There are serious health concerns arising from purchasing the drug without professional advice and regulations, such as buying a fake or expired drug online, through friends, or on the street (Oriakhi et al., 2024).

Determining the exact number of modafinil users is challenging as the drug is used off-label, and the data varies depending on the country and demographics. However, UK-based surveys suggest common use of modafinil, especially for academic study. For example, as early as 2014, The Guardian reported that one in five UK students used modafinil. A recent study using the Reddit platform showed that of 404 respondents, 219 (~54%) used modafinil (Teodorini et al., 2020). Some reports suggest that the use of cognitive enhancers is especially high among medical students (Pighi et al., 2018; Miranda & Borbosa, 2022), which could be due to these individuals being medically informed as well as experiencing intensive academic pressure; however, it is unknown which factors contribute most. Currently, there is no peer-reviewed published data showing the use of modafinil in UK medical schools; however, some studies suggest that the use of other stimulants is common (Plumber et al., 2021; Merwid-Lad et al., 2023).

Despite medical and ethical concerns (Daubner et al., 2021), there is a lack of consensus across academic establishments. Some call for the ban of all “smart drugs” including modafinil, while others advocate for regulated and informed use of modafinil as the safest of the cognitive enhancers. In this case report, we discuss findings from a survey conducted to investigate the awareness, use, and opinions regarding modafinil among medical students. The goal is to identify concerns about modafinil use and leverage these insights to enhance student support services, ultimately providing better assistance to students regarding such drugs.

## Methods

### 1. Participants

Upon approval of the project proposal by the ethics committee, approximately two thousand medical students studying in the academic year 2023-2024 were invited to participate in a short anonymous survey using Microsoft Forms platform (Supplemental information, Appendix I). The invitation emphasised that the survey was anonymous. Before undertaking the survey, the students were required to read the participant information sheet and sign a consent form (Supplemental information, Appendix II), which was accessed via Google Drive, did not require any login details, and contained a description of the project highlighting the potential risks and benefits of participation. All the participants confirmed that they were:

- over 18 years old
- not under influence of psychoactive drugs when undertaking this survey

There was no reward for participation. The responses were collected over 14 weeks (26^th^ February 2024 - 4^th^ June 2024).

### 2. Survey

The survey questionnaire was based on Teodorini et al., 2020 with some minor amendments (Supplemental information, Appendix III). All the questions were related to modafinil, and no demographic data was collected due to a lack of evidence suggesting that demographic factors affect modafinil pharmacodynamics/pharmacokinetics or physiological response in an adult population. The questionnaire was divided into two main sections.

Section I – Modafinil use.

This section included 18 questions covering the awareness of modafinil, its use, frequency of use, dosage taken, motivations for use, perceived risks and benefits, and any perceived long-term effects. All questions had a “prefer not to say” option in case the question is too sensitive to answer.

Section II – Opinion questions. This section contained 8 statements for the participants to express their opinions on the topic (Teodorini et al., 2020) using a scale from strongly agree, agree, neutral, disagree, to strongly disagree.

The final page of the survey included a debrief with a thank you message (Supplemental information, Appendix IV) and a button for submitting the data for analysis.

## Results

### 1. Survey response rate

The invitation was extended to approximately 2,000 students, regardless of their familiarity or experience with modafinil. The survey was anonymous and did not collect any demographic data (e.g., age, gender). Based on previous anonymous surveys conducted by our department at the same time, an expected response rate of 7%-33% had been anticipated (unpublished); therefore, the survey aimed for at least 100 responses to achieve a 10% margin of error at a 95% confidence level. However, after 14 weeks, only 33 responses had been submitted (1.65%). This low response rate makes it challenging to draw conclusive findings and raises questions about the effectiveness of surveys as a tool for investigating drug use among medical students. While the small sample size limits the generalisability of the results, the data gathered still provided a foundation for understanding trends and identifying key factors related to modafinil use among medical students.

### 2. Awareness of Smart Drugs and Modafinil

Approximately 75% of the participants were aware of smart drugs and over half indicated that they would take a drug to increase their attention, concentration, and memory (Figure 1A, B). Interestingly, many of those aware of smart drugs, were not aware of modafinil (45%), suggesting higher familiarity with other cognitive enhancers (Figure 1C). Most participants had heard about smart drugs from their friends (53%) and the media (22%), with very few learning about modafinil from academic literature (7%) or from a medical professional (7%) (Figure 1D, Supplemental Table 1).

**Figure 1.**
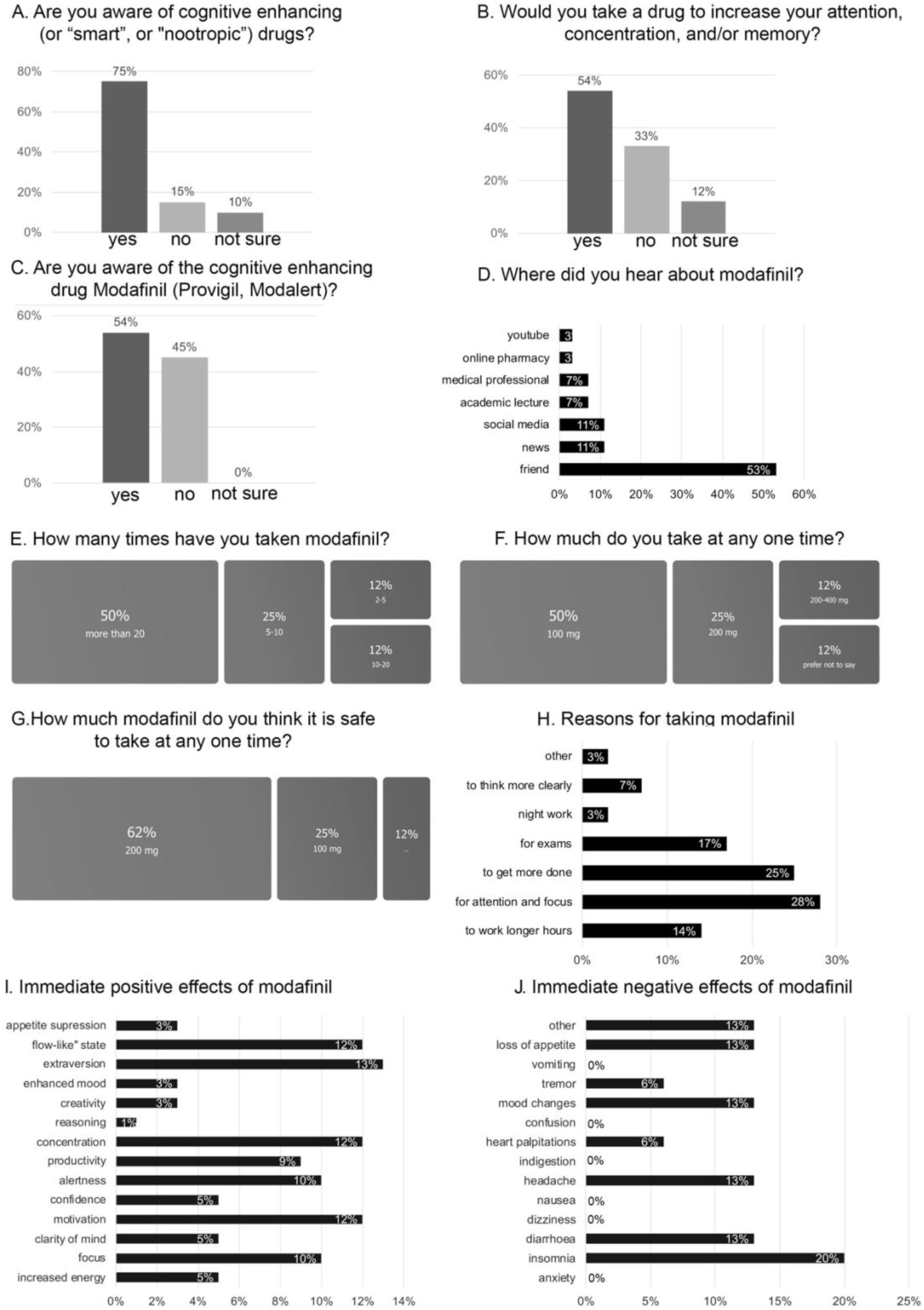
Awareness and use of modafinil in medical students.

### 3. The Use of Modafinil

Of the participants who were aware of modafinil, 44% had used the drug. Half of these users took the drug more than 20 times, three or more times a week. Some respondents reported moderate use, taking the drug about 5-10 times, six times per year or less. The typical dose reported was 100 mg, while more frequent users took doses ranging from 200 mg up to 400 mg (Figure 1E, F, G). Users typically did not combine modafinil with any other drugs, although one respondent reported using it with caffeine.

The reasons for taking modafinil included “for attention and focus” (28%), increased productivity (25%), for exams (17%), and to work longer hours (14%). Other less common reasons included “to think more clearly” and for night work. All users reported the drug as effective, except for one respondent who took 200 mg 3-5 times per week and was unsure if the drug was helpful (Figure 1H).

Immediate positive effects of modafinil included increased motivation, concentration, ability to work in a “flow-like” state, alertness, and focus. Less frequent positive experiences included increased energy, clarity of mind, confidence, creativity, extraversion, and improved mood. Some respondents also experienced appetite suppression, which they viewed positively. Some of these effects were noted to be long-lasting (Figure 1I, J, Supplemental Table 1).

Modafinil was linked to several immediate negative effects, with the most frequent being insomnia, followed by diarrhoea, headache, mood changes, and loss of appetite. Less commonly reported negative effects included tremors, heart palpitations, and fatigue. Some effects, such as insomnia, low mood, and loss of appetite, were reported as long-lasting (Supplemental Table 1). Interestingly, most modafinil users did not feel dependent on the drug, except for one individual who used it more frequently and responded as “not sure” (Supplemental Table 1).

### 4. Opinions about modafinil

The second section of the survey explored opinions about modafinil use for cognitive enhancement, regardless of whether respondents were aware of smart drugs or modafinil, and irrespective of their usage (Figure 2, Supplemental Table 1). Most participants (38%) did not believe they would be endangering themselves by using the drug, while others expressed uncertainty (33%) or considered modafinil potentially risky (27%). Further questions indicated a moderate level of concern about modafinil’s safety, with 45% of participants feeling that it was not entirely safe to take the drug (Figure 2A).

**Figure 2.**
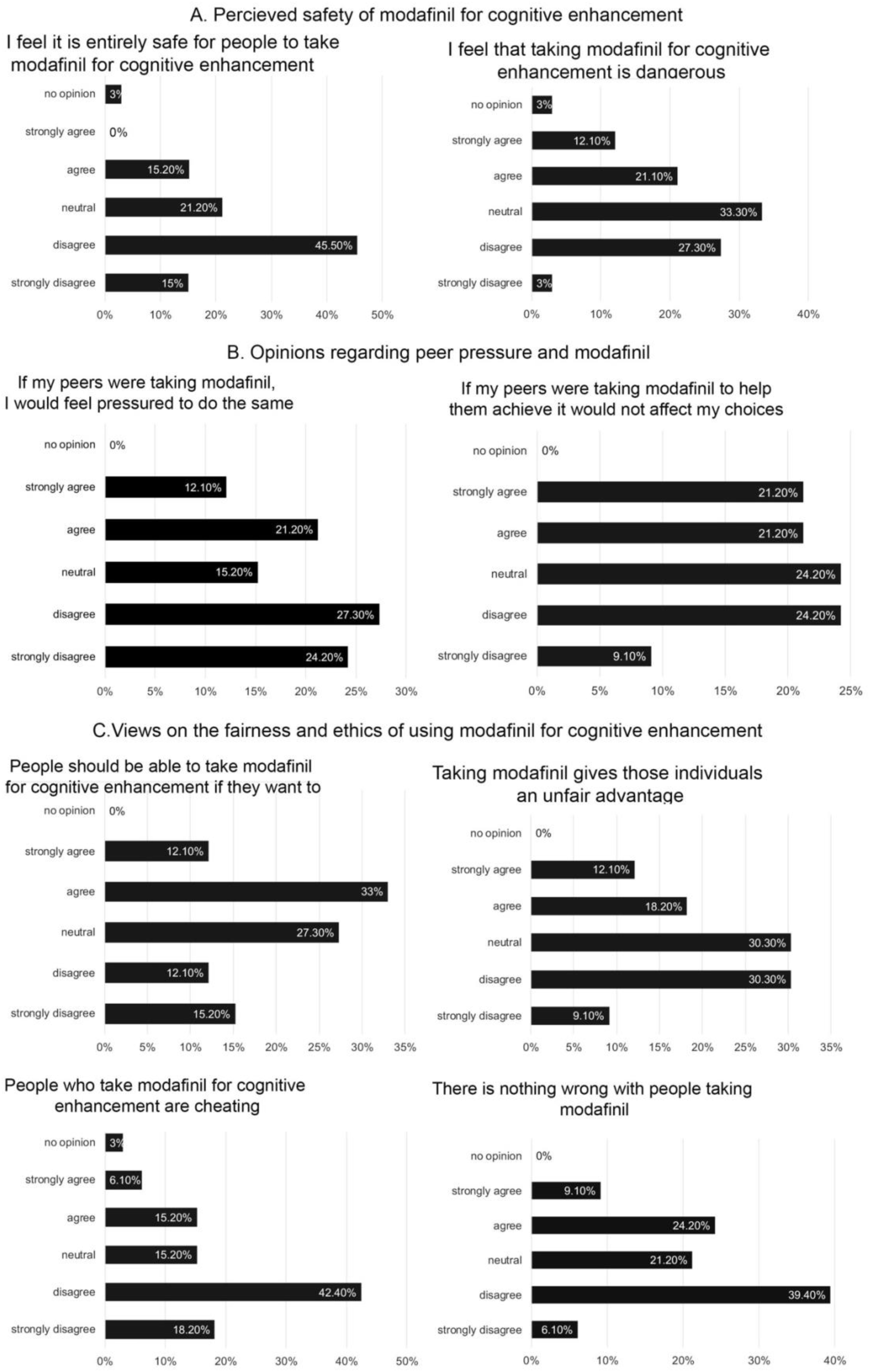
Opinions about the use of modafinil for cofnitive enhancement.

Opinions on peer pressure were mixed: many participants indicated they would not feel pressured to take the drug if their peers were using it for cognitive enhancement, while others admitted their decisions could be influenced (Figure 2B). Furthermore, most respondents did not believe that using modafinil would give users an unfair advantage (Figure 2C). Interestingly, 100% of modafinil users did not consider it cheating, while only 48% of non-users felt the same (24% were neutral or had no opinion, and 28% agreed; Supplemental Table 1). Most participants disagreed with the statement that ‘there is nothing wrong with people taking modafinil,’ although they supported individuals’ freedom to choose to use modafinil for cognitive enhancement if they wished (Figure 2C, Supplemental Table 1).

## Discussion

Online surveys are a popular tool for studying various behaviours, opinions, and trends across diverse populations, including medical students (Braun et al, 2021). They offer several advantages, such as cost-effectiveness, the ability to reach the target audience, and the convenience for respondents to complete the survey at their own pace. However, conducting surveys presents challenges, one of which is low engagement, particularly when surveys do not offer reward for participation (Levefer et al., 2006).

For the survey described in this case report, low response rate was the main limitation that significantly compromised the study. Despite broad invitation and assurance of anonymity as well clarifications that the survey did not imply any illegal activity, the response rate was only 1.65%. This low rate makes it challenging to draw definitive conclusions. It stands in stark contrast to the response rate of other online surveys on different topics, such as learning support, conducted concurrently at our institution (personal communication). This disparity suggests that survey fatigue (Tater et al., 2023) is unlikely to be the sole explanation for the low response rate.

Investigating and addressing why students did not effectively respond to the survey poses challenges. Several factors could be considered, including stigma and the sensitive nature of the topic, which may have made students hesitant to participate. Some students might have perceived questions about drug use as invasive of their privacy, potentially contributing to lower participation rates.

Another significant factor, particularly pertinent in a medical environment, could be fear of repercussions. It’s possible that students distrusted the anonymity and confidentiality of the survey, fearing that their responses could lead raise concerns about their fitness to practice (GMC, UK). In addition to the low response rate, our survey had other limitations inherent to the online survey format, such as response bias and technical issues, which could potentially skew the results.

Despite all the challenges, the data collected can still provide valuable preliminary insights. For instance, our findings highlight that a high awareness (75%) of modafinil and other smart drugs, which may include Aderall^®^ (Amphetamine and Dextroamphetamine), Ritalin^®^ (Methylphenidate), Aricept^®^ (Donepezil), and Namenda^®^ (Memantine). One of the most concerning findings was that the respondents primarily learnt about these drugs from friends and the media rather than from health professionals or academic literature. This trend may be attributed to media portrayal, which often emphasises the potential benefits of these drugs rather than their risks (Partridge et al., 2011).

It is important that individuals make an informed decision based on high-quality information about the risks associated with taking the drug. Therefore, there is a need for an agreed policy and a comprehensive professionally curated advice issued to medical students regarding the use of modafinil and other smart drugs to mitigate the risks.

Modafinil appeared to enhance perceived cognitive performance, with many respondents noting some of the positive effects being long-lasting. They also reported immediate negative side effects, primarily insomnia, which is expected given modafinil’s primary application, i.e. narcolepsy. Importantly, all observed effects of modafinil and side effects reported in our survey have been documented in the literature (Kredlow et al., 2019) and no new effects were reported in the “other” option. With regards to possible dependence on modafinil, none of the respondents considered themselves dependent with only one respondent being unsure. This finding aligns with previous literature suggesting a low addiction potential for modafinil (Myrick et al., 2006). Moreover, modafinil has been proposed for treating addiction to other stimulants such as cocaine (Kapman, 2019). However, there are rare case reports of physical dependence (Krishnan and Chary, 2015; Alacam et al., 2018), highlighting potential dangers, particularly with long-term use of the drug. All these concerns are especially relevant to students, as taking modafinil at a young age can have a long-lasting impact on their future well-being.

With regards to opinions on modafinil, respondents, irrespective of their experience with the drug, expressed willingness to use it for cognitive enhancement and generally harboured moderate concerns about its safety. Interestingly, peer pressure was not a significant factor influencing the decision to use modafinil. Furthermore, respondents did not perceive it as “cheating”, aligning with recent reports (Kesta and Newton, 2024).

In conclusion, our report indicates that investigating smart drug use among medical students needs methodologies beyond online surveys. The sensitive nature of the topic, combined with potential distrust in respondents’ anonymity and concerns about repercussions, may limit the effectiveness of such surveys. Alternative approaches could involve analysing smart drugs and their metabolites within the campus environment during exam periods to assess the prevalence of smart drug use and identify the most commonly used substances among medical students. These insights would facilitate the development of targeted student support strategies aimed at mitigating the risks associated with smart drug use.

## Declaration of conflicting interests

none

## Funding

none

## Ethical approval

Queen Mary Ethics of Research Committee reference number: QME23.0113

All the data is available and is included in the manuscript as main figures and supplemental information

## Data Availability

All data produced in the present work are contained in the manuscript

## Supplemental information

### Appendix I: Study ad

Have you ever heard about “smart drugs’ such as modafinil?

Are you curious about the topic and motivations of Modafinil users?

Join our survey and be part of research on opinions about smart drugs and the use of modafinil!

You don’t need to have taken the drug or any drugs to participate, although anyone who has would contribute greatly! This 10-minute survey is completely confidential and anonymous so no details that could identify you will be recorded. Importantly, we will not ask any questions that would imply any illegal activity.

If you are over 18 years of age and have an opinion about this topic you are most welcome to participate.

### Appendix II: Consent Form

Participant Information Sheet

**Study title**

Awareness and use of the “cognitive enhancer” prescription drug modafinil in medical students

**Researcher’s name […]**

**Ethics of Research Committee reference number:** […]

**Invitation paragraph**

You are being invited to participate in a study that aims to investigate medical students’ awareness, opinions, and off-label use of the prescription drug modafinil, often referred to as a “limitless” or a ‘smart drug’. Before you decide to participate, it is essential that you understand the purpose, procedures, potential risks, and benefits of this study. Please take your time to read this form carefully and discuss with others if you wish. If you would like more information, please contact [researcher’s contact details]

**Background**

‘Smart drugs’, also referred to as ‘cognitive enhancers’ or ‘nootropics’, are prescription drugs claimed to enhance cognitive function. Over the past decade, there has been a notable increase in reports suggesting that healthy people take these drugs off-label in high pressure situations. Modafinil is one such ‘smart drug’ that is prescribed in narcolepsy, sleep apnoea, and shift-worker sleep disorder to promote wakefulness.

Modafinil may offer potential cognitive benefits while being considered well-tolerated with a low risk of addiction; however, it is essential to note that the use carries potential risks, and the long-term effects remain largely unknown. There is also lack of consensus among universities with some proposing bans on the use of all “smart drugs”, including modafinil, while others advocating for the informed and regulated use of modafinil. Therefore, there is a need for further research and informed decision-making regarding the use.

If you would like to read more, click here.

**What is the purpose of the study?**

We aim to assess the awareness, opinions, and experiences of using modafinil among medical students for future initiatives focused on well-being and student support.

**What would taking part involve?**

The study involves an anonymous online questionnaire, which you would need to respond once only and would take about 10 minutes to complete.

**Why am I being invited?**

Your opinions on the topic are invaluable weather it is the first time you hear about modafinil, or you are already aware of this drug, or you may have used it at some point - you can participate regardless.

**Do I have to take part?**

Your participation is entirely voluntary, fully anonymous and you can withdraw from the study at any time. If you find the topic conflicting with your personal beliefs or too invasive, you may not wish to participate.

**What are the possible benefits of taking part?**

Your active participation will help us understand the awareness and the use of modafinil among the students to landscape future projects dedicated to enhancing our student support. Your participation also holds the power to amplify awareness regarding ‘smart drugs’ and their potential side effects. Furthermore, it offers you an important opportunity to introspect your study practices.

**What are the possible disadvantages and risks of taking part?**

Participating in this study carries no known risks. We prioritise your well-being throughout, and it is essential to note that some of the questions may touch upon sensitive topics or experiences. We completely respect your comfort; therefore, you do not have to respond to any questions that you may find uncomfortable or intrusive.

**Expenses and payments** Not applicable

**What information about me will you be collecting?**

None - your participation is fully anonymous, which means we do not collect any personal data, your identity will not be known to the researcher, or anyone involved in the study.

**How will my data be stored and who will have access to it?**

The data will be collected and stored in an anonymous format, the responses will be kept strictly confidential, data will be stored securely and will only be accessible to the researcher. Data will be analysed and reported in aggregate form as opposed to individual responses.

**When and how will my data be destroyed?**

Data will be stored for 5 years and then destroyed.

**How will my data be used and shared?**

The data will form a basis of future studies and will be shared with educators, primarily those involved in student support. Potentially, the results may lead to a publication or become incorporated in future publications.

**Under what legal basis are you collecting this information?**

Please read …privacy notice for research participants containing important information about your personal data and your rights in this respect. If you have any questions relating to data protection, please contact [University’s Data Protection Officer contact details]

**What will happen if I want to withdraw from this study?**

You can withdraw your participation in the study at any time without providing a reason. It will not affect your current or future relationship with the institution or any other organisations in any way.

**What should I do if I have any concerns about this study?**

If you have any concerns about the manner in which the study was conducted, in the first instance, please contact the researcher(s) responsible for the study [Dr…]. If you have a complaint which you feel you cannot discuss with the researchers then you should contact the Research Ethics Facilitators. When contacting the Research Ethics Facilitators, please provide details of the study title, description of the study and reference number (where possible), the researcher(s) involved, and details of the complaint you wish to make.

**Who can I contact if I have any questions about this study?**

**[Research team contact details]**

**Consent form:**

By signing below, I confirm

- I am over 18 years old
- I am not under influence of psychoactive drugs when undertaking this survey
- I have read and understood all of the above information
- I agree to participate

### Appendix III: Questionnaire

#### SECTION I: AWARENESS AND USE

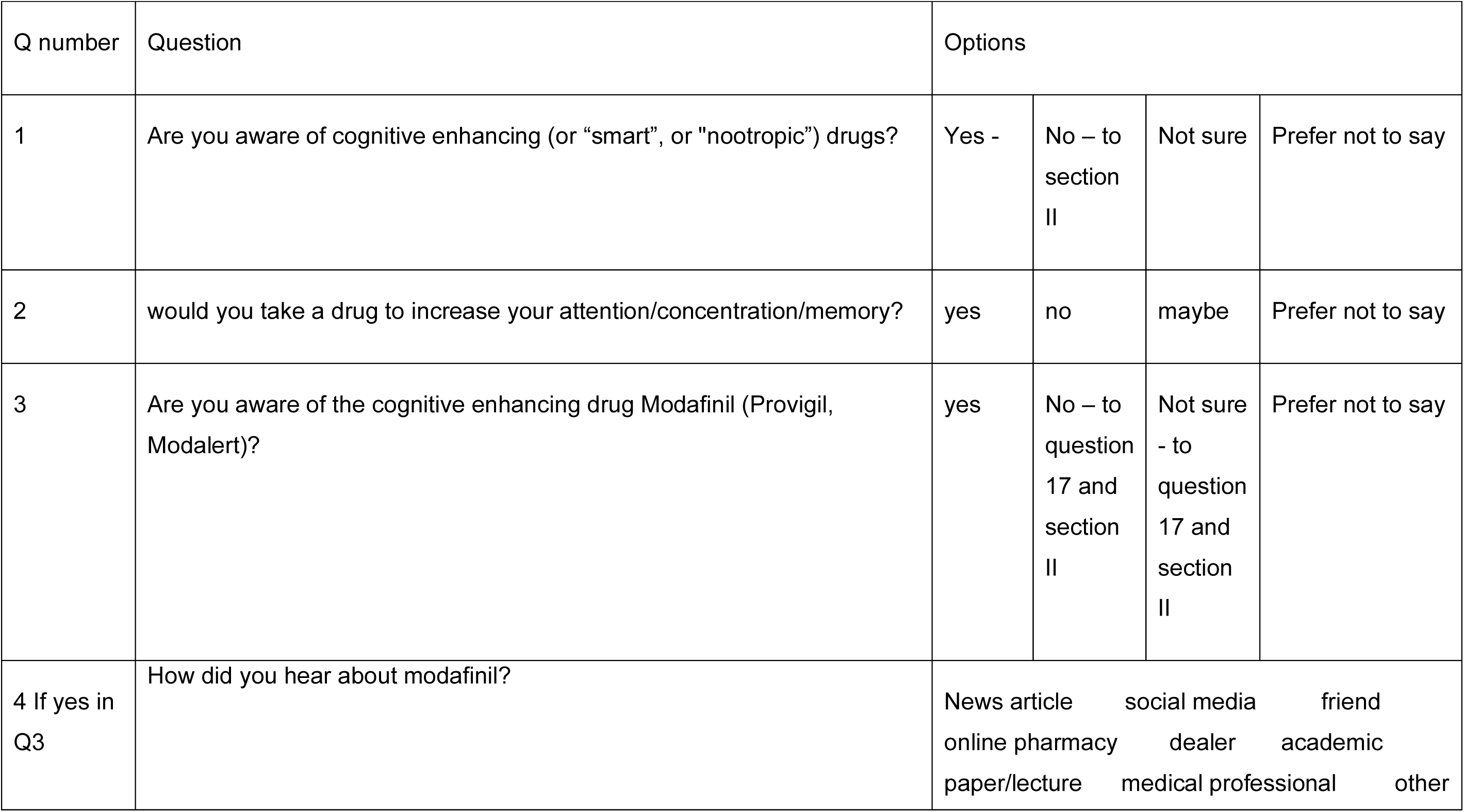
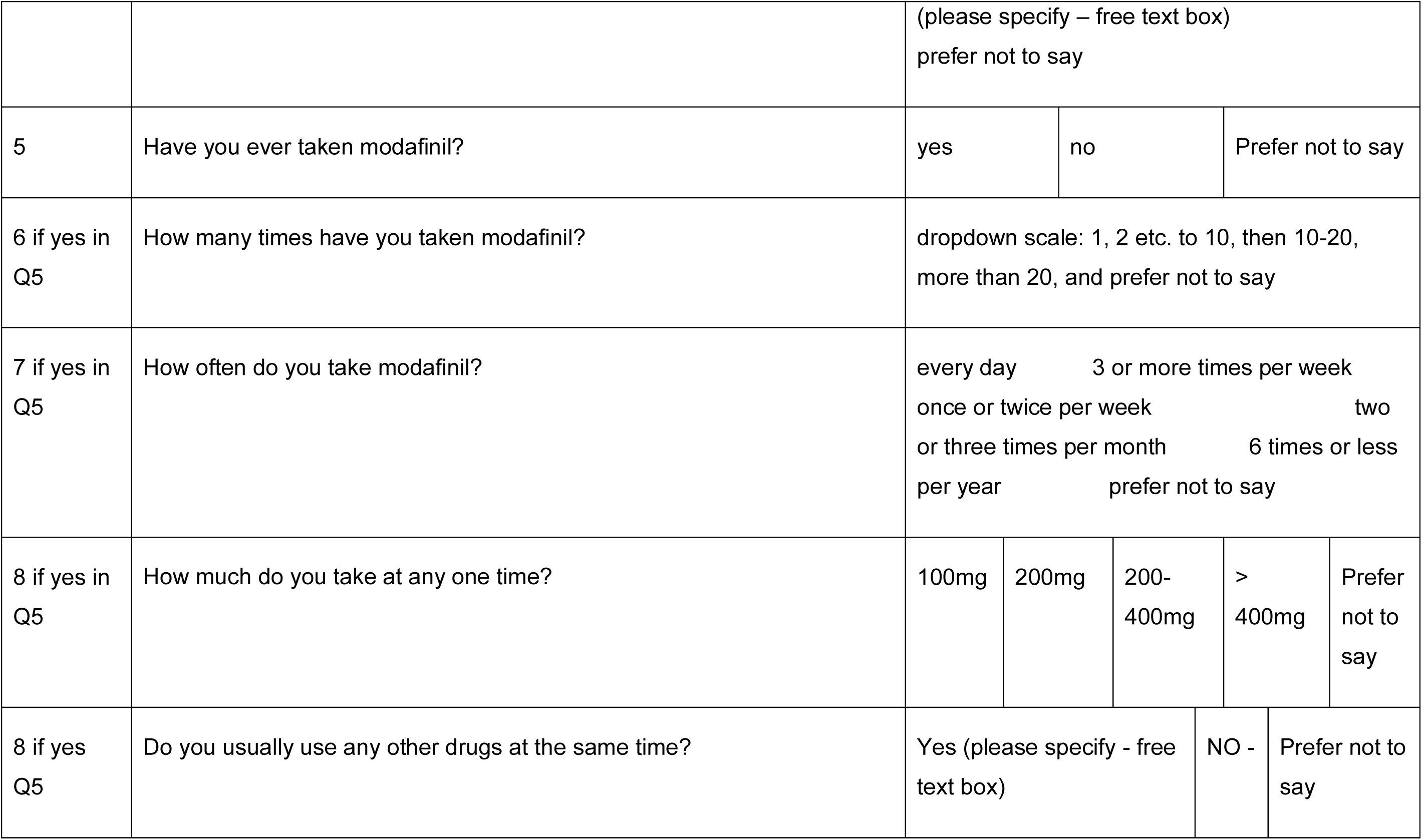
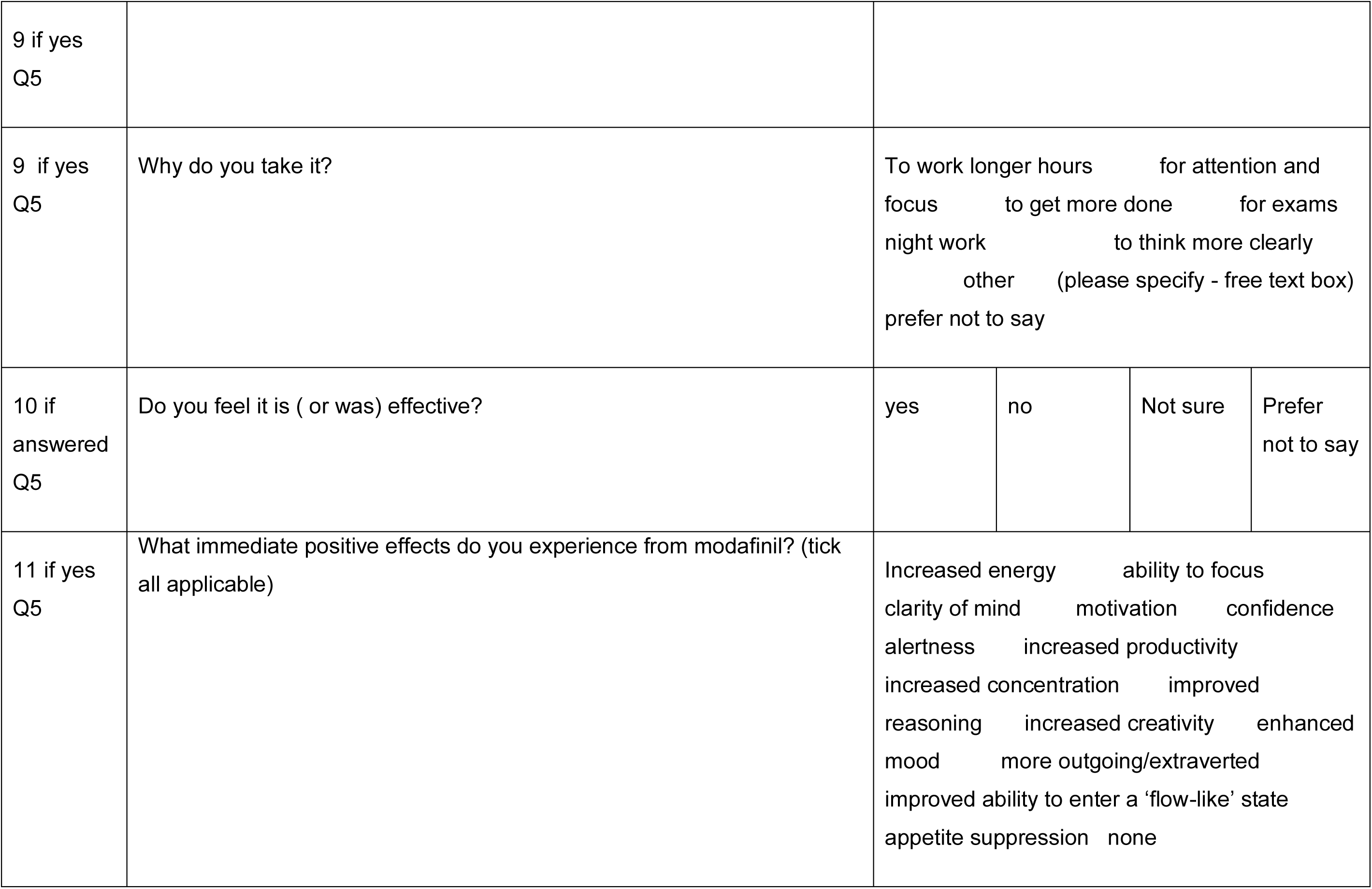
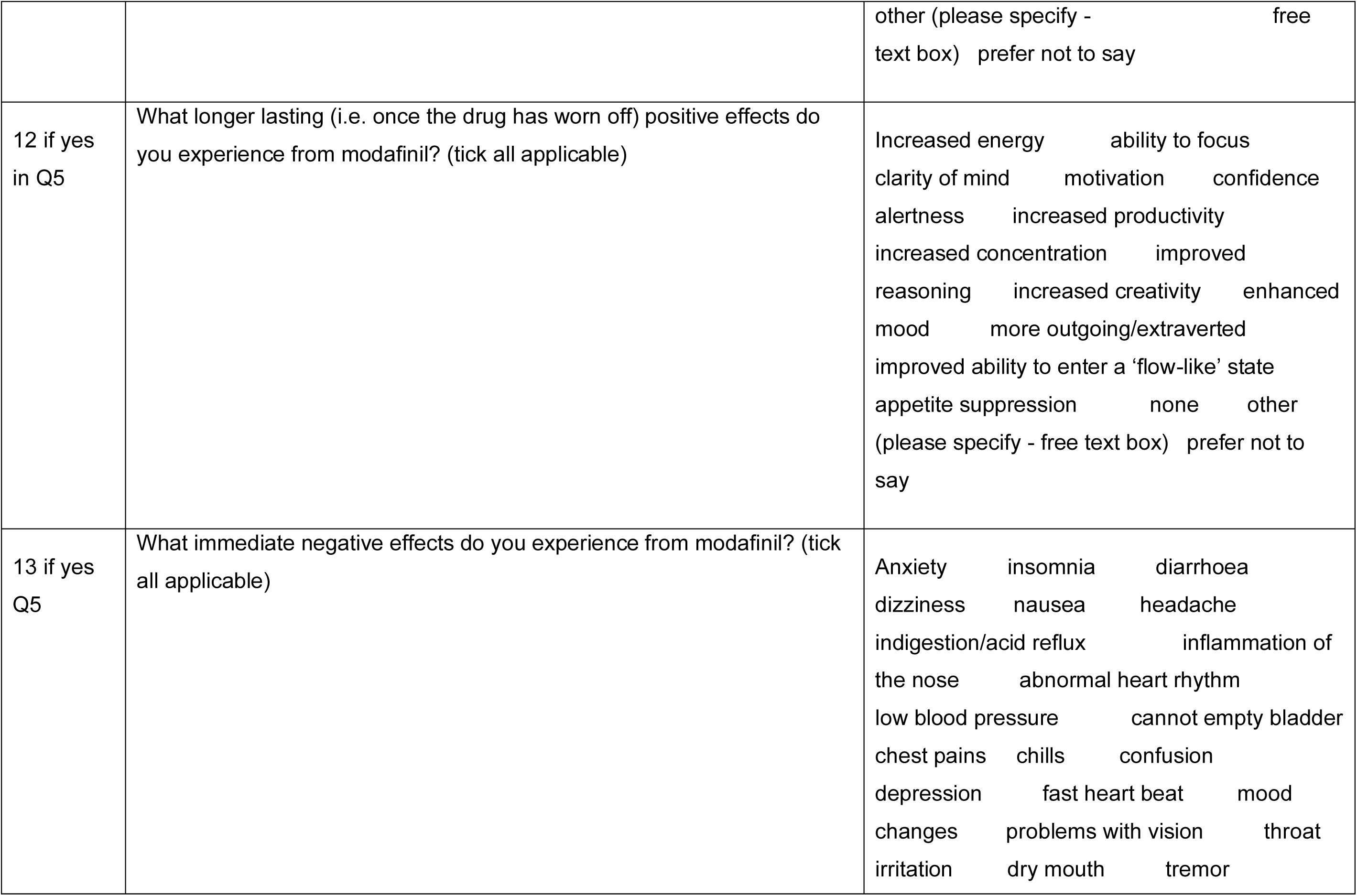
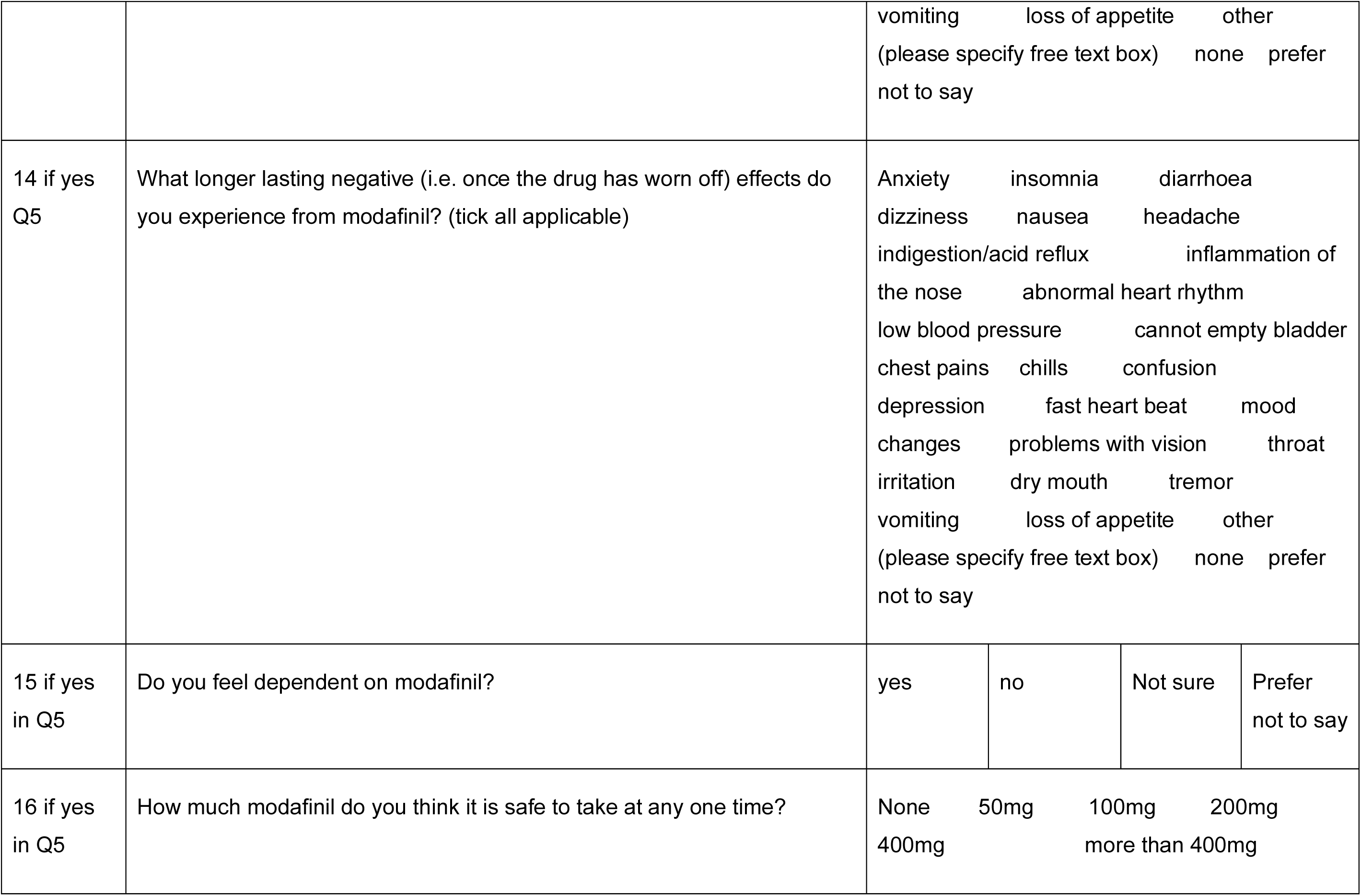
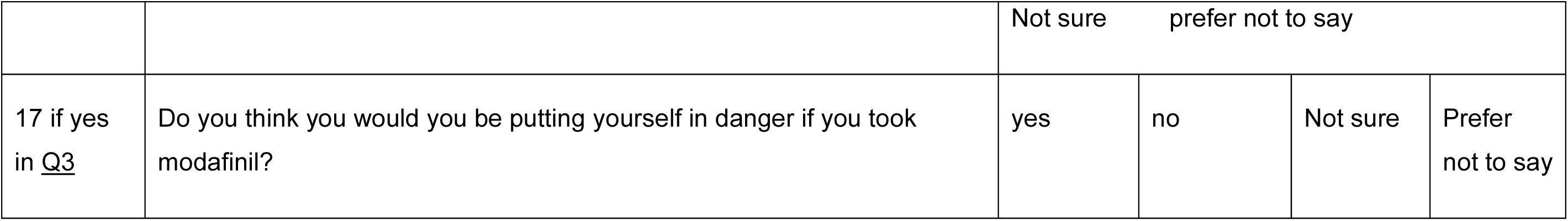

| SECTION I: AWARENESS AND USE |  |  |  |  |  |
| --- | --- | --- | --- | --- | --- |
| Q number | Question | Options |  |  |  |
| 1 | Are you aware of cognitive enhancing (or “smart”, or "nootropic") drugs? | Yes - | No – to section II | Not sure | Prefer not to say |
| 2 | would you take a drug to increase your attention/concentration/memory? | yes | no | maybe | Prefer not to say |
| 3 | Are you aware of the cognitive enhancing drug Modafinil (Provigil, Modalert)? | yes | No – to question 17 and section II | Not sure - to question 17 and section II | Prefer not to say |
| 4 If yes in Q3 | How did you hear about modafinil? | News article<br>social media<br>friend<br>online pharmacy<br>dealer<br>academic<br>paper/lecture<br>medical professional<br>other |  |  |  |

|  |  |  |  |  |  |  |
| --- | --- | --- | --- | --- | --- | --- |
|  |  | (please specify – free text box)<br>prefer not to say |  |  |  |  |
| 5 | Have you ever taken modafinil? | yes | no | Prefer not to say |  |  |
| 6 if yes in Q5 | How many times have you taken modafinil? | dropdown scale: 1, 2 etc. to 10, then 10-20, more than 20, and prefer not to say |  |  |  |  |
| 7 if yes in Q5 | How often do you take modafinil? | every day 3 or more times per week<br>once or twice per week two<br>or three times per month 6 times or less<br>per year prefer not to say |  |  |  |  |
| 8 if yes in Q5 | How much do you take at any one time? | 100mg | 200mg | 200-400mg | > 400mg | Prefer not to say |
| 8 if yes Q5 | Do you usually use any other drugs at the same time? | Yes (please specify - free text box) |  |  | NO - | Prefer not to say |
| 9 if yes<br>Q5 |  |  |  |  |  |
| 9 if yes<br>Q5 | Why do you take it? | To work longer hours for attention and<br>focus to get more done for exams<br>night work to think more clearly<br>other (please specify - free text box)<br>prefer not to say |  |  |  |
| 10 if<br>answered<br>Q5 | Do you feel it is ( or was) effective? | yes | no | Not sure | Prefer<br>not to say |
| 11 if yes<br>Q5 | What immediate positive effects do you experience from modafinil? (tick all applicable) | Increased energy ability to focus<br>clarity of mind motivation confidence<br>alertness increased productivity<br>increased concentration improved<br>reasoning increased creativity enhanced<br>mood more outgoing/extraverted<br>improved ability to enter a 'flow-like' state<br>appetite suppression none |  |  |  |

|  |  |  |
| --- | --- | --- |
|  |  | other (please specify - free text box) prefer not to say |
| 12 if yes in Q5 | What longer lasting (i.e. once the drug has worn off) positive effects do you experience from modafinil? (tick all applicable) | Increased energy ability to focus<br>clarity of mind motivation confidence<br>alertness increased productivity<br>increased concentration improved reasoning increased creativity enhanced mood more outgoing/extraverted<br>improved ability to enter a 'flow-like' state<br>appetite suppression none other (please specify - free text box) prefer not to say |
| 13 if yes Q5 | What immediate negative effects do you experience from modafinil? (tick all applicable) | Anxiety insomnia diarrhoea<br>dizziness nausea headache<br>indigestion/acid reflux inflammation of the nose abnormal heart rhythm<br>low blood pressure cannot empty bladder<br>chest pains chills confusion<br>depression fast heart beat mood changes problems with vision throat irritation dry mouth tremor |
|  |  | vomiting loss of appetite other<br>(please specify free text box) none prefer<br>not to say |  |  |  |
| 14 if yes<br>Q5 | What longer lasting negative (i.e. once the drug has worn off) effects do you experience from modafinil? (tick all applicable) | Anxiety insomnia diarrhoea<br>dizziness nausea headache<br>indigestion/acid reflux inflammation of<br>the nose abnormal heart rhythm<br>low blood pressure cannot empty bladder<br>chest pains chills confusion<br>depression fast heart beat mood<br>changes problems with vision throat<br>irritation dry mouth tremor<br>vomiting loss of appetite other<br>(please specify free text box) none prefer<br>not to say |  |  |  |
| 15 if yes<br>in Q5 | Do you feel dependent on modafinil? | yes | no | Not sure | Prefer<br>not to say |
| 16 if yes<br>in Q5 | How much modafinil do you think it is safe to take at any one time? | None 50mg 100mg 200mg<br>400mg more than 400mg |  |  |  |

|  |  | Not sure prefer not to say |  |  |  |
| --- | --- | --- | --- | --- | --- |
| 17 if yes<br>in <u>Q3</u> | Do you think you would you be putting yourself in danger if you took modafinil? | yes | no | Not sure | Prefer<br>not to say |

#### SECTION II: OPINION QUESTIONS

ALL HAVE THE FOLLOWING ANSWER FORMAT

*strongly disagree disagree neutral agree strongly agree prefer not to say no opinion*

I feel it is entirely safe for people to take modafinil for cognitive enhancement

I feel that taking modafinil for cognitive enhancement is dangerous

If my peers were taking modafinil to help them achieve more I would feel pressured to do the same

If my peers were taking modafinil to help them achieve it would not affect my choices

People should be able to take modafinil for cognitive enhancement if they want to

Taking modafinil gives those individuals an unfair advantage

People who take modafinil for cognitive enhancement are cheating

There is nothing wrong with people taking modafinil

### Appendix IV: Debrief

We sincerely thank you for your participation in our study. Your contributions are invaluable to us, and we are grateful for your time and effort. Please rest assured that your responses are fully anonymous and confidential, ensuring your privacy.

Our study aims to gather important data to enhance our student support and guidance programs. The analysed combined data will be shared with the student support services and your input will directly contribute to our excellence in student support mission.

If you have any concerns or questions about modafinil, we strongly advise consulting with your healthcare professional for guidance. Furthermore, you can choose to use other resources where you can get a confidential advice:

- FRANK®: Telephone: 0300 1236600 or visit https://www.talktofrank.com/
- Humankind ® https://humankindcharity.org.uk/drug-and-alcohol-recovery/#:~:text=Humankind%20provides%20integrated%20substance%20misuse,both%20adults%20and%20young%20people.
- Barts and The London Student Association Welfare https://www.bartslondon.com/welfare

Additionally, if you would like to access MHRA UK advice regarding the safe use of Modafinil, you can find more information by clicking on the links below.

- https://www.gov.uk/government/news/freshers-warned-to-be-smart-and-avoid-modafinil
- https://www.gov.uk/drug-safety-update/modafinil-provigil-increased-risk-of-congenital-malformations-if-used-during-pregnancy
- https://www.gov.uk/drug-safety-update/modafinil-provigil-now-restricted-to-narcolepsy

Once again, thank you for your participation, and we appreciate your commitment to helping us improve our student support services

## Notes

### Competing Interest Statement

The authors have declared no competing interest.

### Funding Statement

This study did not receive any funding

### Author Declarations

Queen Mary Ethics of Research Committee reference number: QME23.0113

