## Supplemental Table 1 for "Awareness and use of the “cognitive enhancer” prescription drug modafinil in medical students"

| Id | Email | Name | By ticking the boxes belo |
|---|---|---|---|
| 2 | anonymous | | I am over 18 years old;I a |
| 3 | anonymous | | I am over 18 years old;I a |
| 4 | anonymous | | I am over 18 years old;I a |
| 5 | anonymous | | I am over 18 years old;I a |
| 6 | anonymous | | I am over 18 years old;I a |
| 7 | anonymous | | I am over 18 years old;I a |
| 8 | anonymous | | I am over 18 years old;I a |
| 9 | anonymous | | I am over 18 years old;I a |
| 10 | anonymous | | I am over 18 years old;I a |
| 11 | anonymous | | I am not under influence |
| 12 | anonymous | | I am over 18 years old;I a |
| 13 | anonymous | | I am over 18 years old;I a |
| 14 | anonymous | | I am over 18 years old;I a |
| 15 | anonymous | | I am over 18 years old;I a |
| 16 | anonymous | | I am over 18 years old;I a |
| 17 | anonymous | | I am not under influence |
| 18 | anonymous | | I am over 18 years old;I a |
| 19 | anonymous | | I am over 18 years old;I a |
| 20 | anonymous | | I am over 18 years old;I h |
| 21 | anonymous | | I am over 18 years old;I a |
| 22 | anonymous | | I am over 18 years old;I a |
| 23 | anonymous | | I am over 18 years old;I a |
| 24 | anonymous | | I am over 18 years old;I a |
| 25 | anonymous | | I am over 18 years old;I a |
| 26 | anonymous | | I am over 18 years old;I h |
| 27 | anonymous | | I am over 18 years old;I a |
| 28 | anonymous | | I am over 18 years old;I a |
| 29 | anonymous | | I am over 18 years old;I a |
| 30 | anonymous | | I am over 18 years old;I a |
| 31 | anonymous | | I am over 18 years old;I a |
| 32 | anonymous | | I am over 18 years old;I a |
| 33 | anonymous | | I am over 18 years old;I a |
| 34 | anonymous | | I am over 18 years old;I a |

| Are you aware of cogniti | Would you take a drug t | Are you aware of the co | How did you hear about |
|---|---|---|---|
| Yes | No | Yes | social media; |
| Yes | No | Yes | friend; |
| Yes | Yes | Yes | news article;friend; |
| Not sure | No | No | |
| Yes | Yes | Yes | friend;social media; |
| Yes | Yes | Yes | YouTube ; |
| Yes | Yes | No | |
| Yes | Yes | Yes | friend; |
| Yes | Yes | No | |
| Not sure | Yes | No | |
| Yes | No | No | |
| Yes | Not sure | No | |
| Yes | Yes | Yes | friend;academic paper/le |
| No | Yes | No | |
| Yes | Yes | Yes | friend; |
| Not sure | Yes | No | |
| Yes | Yes | Yes | friend; |
| Not sure | Not sure | Yes | medical professional; |
| No | Yes | No | |
| No | No | No | |
| Yes | No | Yes | friend; |
| Yes | No | Yes | news article;friend; |
| Yes | No | No | |
| Yes | No | No | |
| Yes | Not sure | No | |
| Yes | Yes | Yes | friend;medical professio |
| Yes | No | Yes | social media;news article |
| Yes | Not sure | Yes | friend; |
| Yes | Yes | No | |
| Yes | No | Yes | friend; |
| Not sure | Yes | No | |
| Yes | Yes | Yes | friend; |
| Yes | Yes | Yes | friend; |

| Have you ever taken mo | How many times have y | How often do you take r | How much do you take a |
|---|---|---|---|
| No | | | |
| No | | | |
| Yes | more than 20 | six times or less per year | 100 mg |
| No | | | |
| No | | | |
| Yes | more than 20 | once or twice per week | 100 mg |
| Yes | 5-10 | three or more per week | 200 mg |
| Yes | 2-5 | six times or less per year | 200-400 mg |
| Yes | 5-10 | prefer not to say | prefer not to say |
| No | | | |
| No | | | |
| No | | | |
| Yes | more than 20 | three or more per week | 100 mg |
| No | | | |
| No | | | |
| No | | | |
| Yes | 10-20 | two or three times a mo | 200 mg |
| Yes | more than 20 | three or more per week | 100 mg |

| Do you usually use any c | Why do you take modafi | Do you feel taking moda | What immediate positiv |
|---|---|---|---|
| Yes - please specify in th | for attention and focus;t | Yes | Increased energy;Ability |
| No; | to work longer hours;for | Yes | Ability to focus;Motivatic |
| No; | for attention and focus;S | Not sure | Alertness;Appetite supre |
| No; | for attention and focus;t | Yes | Motivation;Alertness;Inc |
| No; | for attention and focus;t | Yes | Ability to focus;Motivatic |
| No; | to work longer hours;for | Yes | Ability to focus;Motivatic |
| No; | to work longer hours;for | Yes | Ability to focus;Clarity of |
| No; | to work longer hours;for | Yes | Increased energy;Ability |

| What longer lasting posi | What immediate negativ | What longer lasting nega | Do you feel dependent c |
|---|---|---|---|
| Increased concentration; | Headache;Diarrhoea; | Insomnia; | No |
| Improved ability to work | Insomnia; | | No |
| ssion; | mood changes;loss of ap | Possible dependence; | Not sure |
| Increased energy;More c | Diarrhoea;abnormal hea | Headache;loss of appetit | No |
| on;Clarity of mind;Alertness;Increased concentration;Improved ability to wo | | | No |
| There isn't any; | None ; | depression;mood change | No |
| Confidence; | Insomnia;Headache; | Insomnia; | No |
| Once the medication has | Insomnia;mood changes, | fatigue; | No |

| How much modafinil do | Do you think you would | How do you feel about t | How do you feel about t |
|---|---|---|---|
| | Yes | Neutral | Neutral |
| | Not sure | Strongly Disagree | Strongly agree |
| 200 mg | Yes | Agree | Strongly Disagree |
| | | Neutral | Agree |
| | No | Disagree | Disagree |
| | No | Neutral | Disagree |
| | | Strongly Disagree | Agree |
| 200 mg | No | Neutral | Neutral |
| | | No opinion | No opinion |
| | | Agree | Disagree |
| | | Strongly Disagree | Strongly agree |
| | | Disagree | Neutral |
| 200 mg | Not sure | Disagree | Neutral |
| | | Neutral | Disagree |
| 100 mg | Not sure | Disagree | Neutral |
| | | Disagree | Neutral |
| not sure | Yes | Disagree | Agree |
| | Not sure | Neutral | Neutral |
| | | Agree | Neutral |
| | | Disagree | Agree |
| | Yes | Strongly Disagree | Strongly agree |
| | Not sure | Disagree | Strongly agree |
| | | Strongly Disagree | Agree |
| | | Disagree | Neutral |
| | | Disagree | Disagree |
| 100 mg | No | Agree | Disagree |
| | Yes | Disagree | Agree |
| | Not sure | Disagree | Disagree |
| | | Disagree | Neutral |
| | No | Disagree | Agree |
| | | Neutral | Neutral |
| 200 mg | No | Disagree | Disagree |
| 200 mg | No | Agree | Disagree |

| What do you think of th | What do you think of th | How do these statement | How do these statement |
|---|---|---|---|
| Neutral | Neutral | Neutral | Neutral |
| strongly disagree | Strongly agree | Disagree | Neutral |
| strongly disagree | Strongly agree | Strongly agree | strongly disagree |
| Disagree | agree | agree | agree |
| agree | Disagree | agree | Disagree |
| Disagree | agree | agree | Disagree |
| Strongly agree | strongly disagree | strongly disagree | Strongly agree |
| Disagree | Neutral | agree | Disagree |
| Disagree | agree | agree | Disagree |
| Disagree | agree | agree | Disagree |
| strongly disagree | Strongly agree | strongly disagree | Neutral |
| agree | Disagree | Disagree | Strongly agree |
| strongly disagree | Strongly agree | Neutral | Neutral |
| Strongly agree | strongly disagree | Strongly agree | strongly disagree |
| Neutral | Disagree | Neutral | Disagree |
| Strongly agree | Disagree | Neutral | Strongly agree |
| Disagree | Neutral | agree | Neutral |
| strongly disagree | strongly disagree | agree | Neutral |
| Strongly agree | Neutral | Neutral | agree |
| Neutral | agree | Disagree | agree |
| Disagree | agree | agree | Disagree |
| strongly disagree | Strongly agree | strongly disagree | Disagree |
| strongly disagree | Strongly agree | strongly disagree | agree |
| Disagree | Neutral | Disagree | agree |
| Neutral | Disagree | agree | strongly disagree |
| Neutral | Neutral | Strongly agree | Neutral |
| strongly disagree | Strongly agree | Neutral | Neutral |
| agree | Disagree | strongly disagree | Strongly agree |
| agree | Disagree | Neutral | agree |
| Disagree | agree | Neutral | Disagree |
| agree | Neutral | Neutral | Disagree |
| agree | Neutral | Strongly agree | Neutral |
| agree | Disagree | agree | Neutral |

| What is your opinion on | What is your opinion on |
|---|---|
| Neutral | Neutral |
| No opinion | Disagree |
| strongly disagree | Strongly agree |
| Neutral | agree |
| Disagree | Disagree |
| Disagree | agree |
| Strongly agree | strongly disagree |
| strongly disagree | agree |
| Disagree | agree |
| strongly disagree | agree |
| Neutral | Disagree |
| agree | Disagree |
| Disagree | Disagree |
| strongly disagree | Strongly agree |
| Disagree | agree |
| agree | Neutral |
| Disagree | Neutral |
| Neutral | Neutral |
| Disagree | Neutral |
| agree | Neutral |
| Disagree | Disagree |
| agree | Disagree |
| Strongly agree | strongly disagree |
| Neutral | Disagree |
| strongly disagree | agree |
| strongly disagree | Strongly agree |
| agree | Disagree |
| Disagree | Disagree |
| Disagree | Disagree |
| Disagree | Disagree |
| Disagree | Disagree |
| Disagree | Neutral |
| Disagree | agree |

people taking modafinil
